# Estimating the effectiveness of the Pfizer COVID-19 BNT162b2 vaccine after a single dose. A reanalysis of a study of ‘real-world’ vaccination outcomes from Israel

**DOI:** 10.1101/2021.02.01.21250957

**Authors:** Paul R Hunter, Julii Brainard

**Affiliations:** The Norwich Medical School, University of East Anglia, Norwich, NR4 7TJ, UK

**Author notes:** Corresponding author: Professor Paul Hunter. **Declarations**. Conflict of interest The authors declare that we have no conflict of interest. **Funding** Professor Hunter and Dr. Brainard were funded by the National Institute for Health Research Health Protection Research Unit (NIHR HPRU) in Emergency Preparedness and Response at King’s College London in partnership with Public Health England (PHE) and collaboration with the University of East Anglia. The views expressed are those of the author(s) and not necessarily those of the NHS, the NIHR, UEA, the Department of Health or PHE.

**Keywords:** COVID-19, vaccination, Israel, vaccine effectiveness

## Abstract

A distinctive feature of the roll out of vaccination against SARS-CoV-2 virus in the UK was the decision to delay the timing of the second injection till 12 weeks after the first. The logic behind this is to protect more people sooner and so reduce the total number of severe infections, hospitalisations, and deaths. This decision caused criticism from some quarters due in part to a belief that a single injection may not give adequate immunity. A recent paper based on Israel’s experience of vaccination suggested that a single dose may not provide adequate protection. Here we extract the primary data from the Israeli paper and then estimate the incidence per day for each day after the first injection and also estimate vaccine effectiveness for each day from day 13 to day 24. We used a pooled estimate of the daily incidence rate during days 1 to 12 as the counterfactual estimate of incidence without disease and estimated confidence intervals using Monte Carlo modelling. After initial injection case numbers increased to day 8 before declining to low levels by day 21. Estimated vaccine effectiveness was pretty much 0 at day 14 but then rose to about 90% at day 21 before levelling off. The cause of the initial surge in infection risk is unknown but may be related to people being less cautious about maintaining protective behaviours as soon as they have the injection. What our analysis shows is that a single dose of vaccine is highly protective, although it can take up to 21 days to achieve this. The early results coming from Israel support the UK policy of extending the gap between doses by showing that a single dose can give a high level of protection.

## Introduction

The current pandemic of SARS-CoV-2 is one of the greatest public health challenges to have faced human society for decades. At the time of writing 1^st^ February 2021, over 100 million cases of infection have been reported and over 2 million deaths. Fortunately, the increasing availability of effective vaccines is likely to substantially reduce the future disease burden from this infection. Even so, the substantial ongoing surge in cases and fatalities being seen across many countries means that the rollout of vaccination will come too late for many people.

In order to save lives, the UK Joint Committee on Vaccination and Immunization decided to make the bold recommendation that the gap between first and second doses of COVID vaccinations (DHSC 2021). The recommendation was that rather than give the second dose after days (as was done in the phase 3 trial (Polack et al. 2020)), the second dose should be delayed till 12 weeks after the first dose. The basis of this decision was that although two injections would give better protection to an individual than would one inoculation, giving twice as many people a single injection as soon as possible was likely to reduce the incidence of severe disease in the most individuals. But whilst there was good evidence that extending the gap between doses of the Oxford AstraZeneca vaccine would not adversely affect its ultimate protective effect, no such data were available for the Pfizer-made BNT162b2 mRNA vaccine (Polack et al. 2020; Voysey et al. 2020). It is fair to say that the decision to deviate from the inoculation interval tested in the experimental Pfizer trial prompted much discussion (Sewell et al. 2020). Part of the criticisms were due to the use of estimates of efficacy after a single dose included data from very soon after the injection and therefore before it was reasonable to expect any effect of vaccination. When analysis was restricted to the period when it was reasonable to expect a benefit (ie after 12 days), efficacy estimates were much higher for the Pfizer product (Hunter 2020).

In a pre-print report (Chodick et al. 2021), data were presented on the roll out of the Pfizer vaccine in Israel. In that paper the authors conducted a retrospective cohort study using looking at reported infections in over 500,000 people who had been given the Pfizer vaccine from the day after first immunization to day 24 (Chodick et al. 2021). They estimated that the effectiveness of the Pfizer vaccine prior to the second dose by looking at the incidence of infections during the 13 to 24-day period and compared this with the incidence during the 1 to 12-day period. The authors concluded that the effectiveness of a single dose of the Pfizer vaccine during this 13 to 24-day compared to the 1 to 12-day period was just 51%. However, the authors also noted that in this study that the rate of increase in the cumulative incidence only started to decline after day 18. Despite knowing that for at least five days of their follow-up period there was no apparent benefit from the single dose, they did not then attempt to estimate effectiveness for the later period (after day18) when the vaccine did appear to have an effect. This would then give a better indication of how effective a single dose of the vaccine could be if the second dose was delayed up to 12 weeks.

Given the very large number of people whose records were included in the analysis, it was feasible for us to examine the daily change in vaccine effectiveness and so give a better estimate of the potential effectiveness achievable by a single dose. In this paper we reanalyse the date contained in the paper by Chodick and colleagues in order the determine the potential effectiveness achievable by a single dose of the Pfizer vaccine in a real word situation.

## Methods

The data contained in this analysis were taken from Figure 1 in the pre-print by Chodick and colleagues (2021) by measuring the reported cumulative incidence for each day from Figure 1 using Microsoft Publisher™. From the table of cumulative incidence created from the original paper, the daily incidence was calculated as the change in cumulative incidence over the previous day. As was done by Chodick et al. (2021), we assumed that no effect of the vaccine was observable from day 1 to day 12 and the pooled mean incidence for this period taken to be the incidence without vaccine.

**Figure 1.**
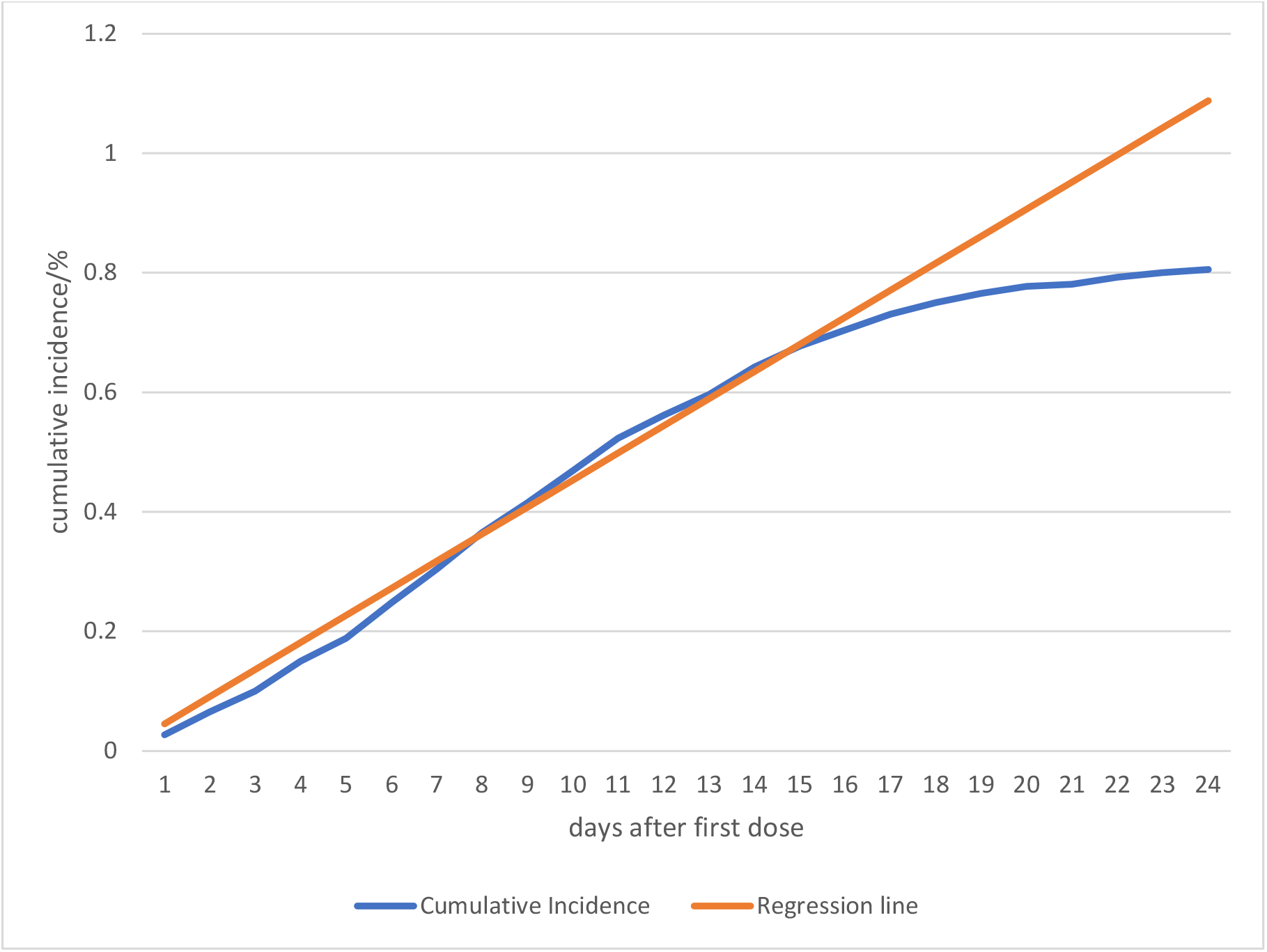
Cumulative incidence (%) compared to extended linear prediction based on incidence from 1st to 12th day after first injection.

We calculated vaccine effectiveness for each day from day 13 to day 24, using the following formula:

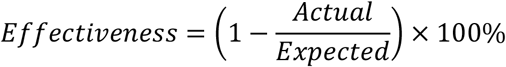

Where *Actual* was the number of cases obtained by multiplying the incidence by numbers at risk for that day and *Expected* was obtained by multiplying the pooled incidence for days 1 to 12 by the numbers at risk for each of the days 13 to 24. The pooled incidence for days 1 to 12 were calculated using the meta-analysis function for proportions within StatsDirect™ (https://www.statsdirect.co.uk/). For both *Actual* and *Expected*, the number of people at risk was the same for each day.

Daily effectiveness was estimated using a Monte Carlo simulation within @Risk™ (https://www.palisade.com/risk/default.asp). Both Actual and Expected case numbers were randomly estimated 10,000 times using the Binomial function. This was recalculated 10,000 times for each day and mean values along with 5^th^ and 95^th^ percentiles, obtained.

## Results

The overall number of actual cases across all 24 days in out model was 3077 which compares closely to 3098 incident cases as reported by Chodick et al. (2021). The small discrepancy is most likely explained by measurement imprecision when estimating numbers from a published graph.

Figure 1 shows the cumulative incidence by day after the first dose with a straight line superimposed derived by extending the regression line based on a linear regression from day 1 to day 12. It can be seen that the actual line follows the regression line closely till about day 14 after which it gradually falls below the straight line. Figure 2 shows the estimated daily incidence. Surprisingly, daily incidence increases strongly after vaccination till about day 8, approximately doubling. Whilst it is not possible to know for certain why this may be the case, there have been concerns that people may believe they are protected as soon as they have had or (indeed have scheduled) their first injection and so start engaging in risky behaviour more than previously (SPI-B 2020). In the Israeli dataset, after day 8, the daily incidence starts to decline substantially till day 21 when in the few remaining days, it seems to level out.

**Figure 2.**
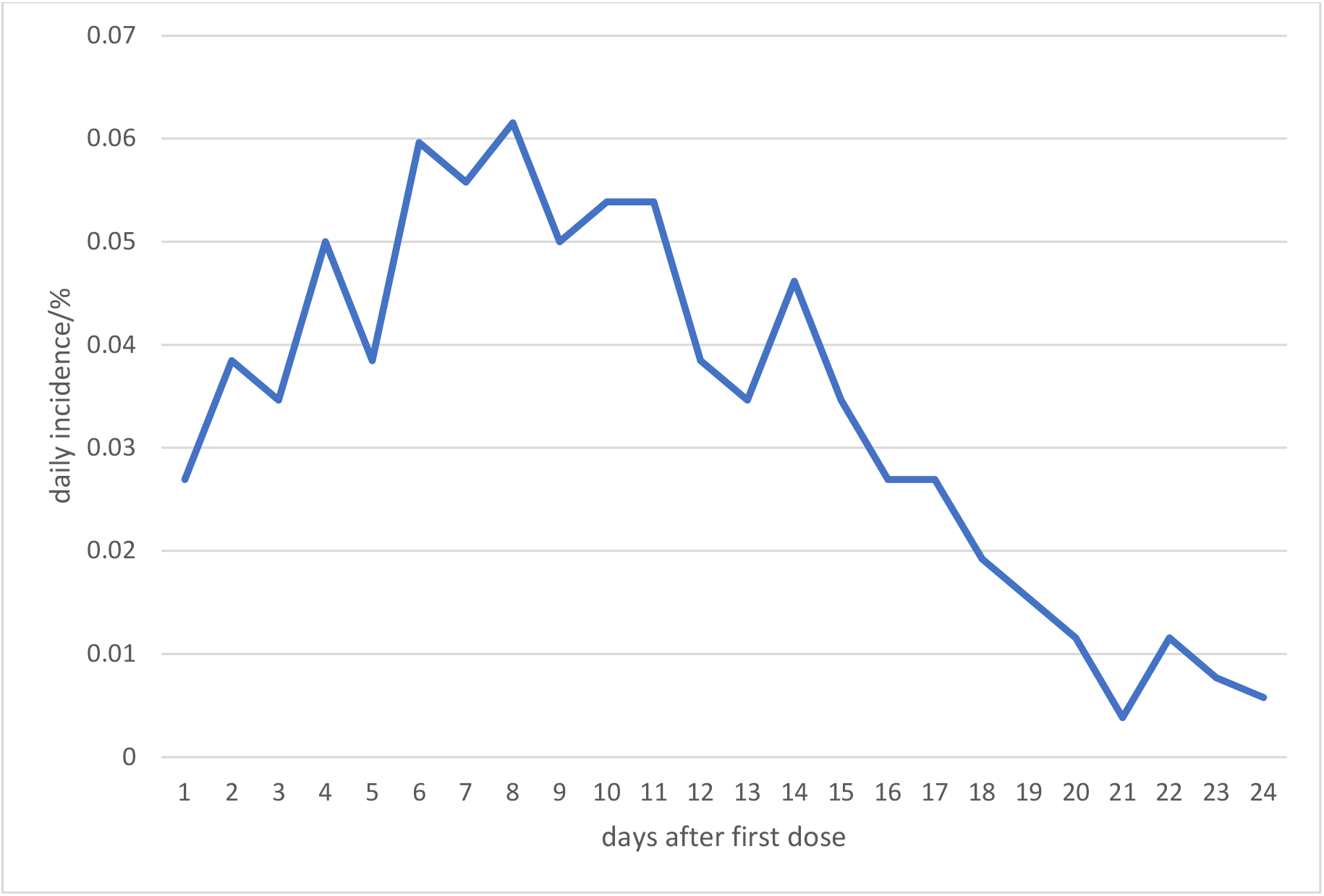
Daily incidence of new infections by days from first dose

Figure 3 shows the estimated effectiveness of the Pfizer vaccine on each day from 13 to 24 days after the first injection. It can be seen that the at day 14 there was no apparent effect of the vaccine but from then on till day 21 the effectiveness reached 91% (90% credible intervals: 83 to 98%). After then the effectiveness levelled off, and case numbers became quite low.

**Figure 3.**
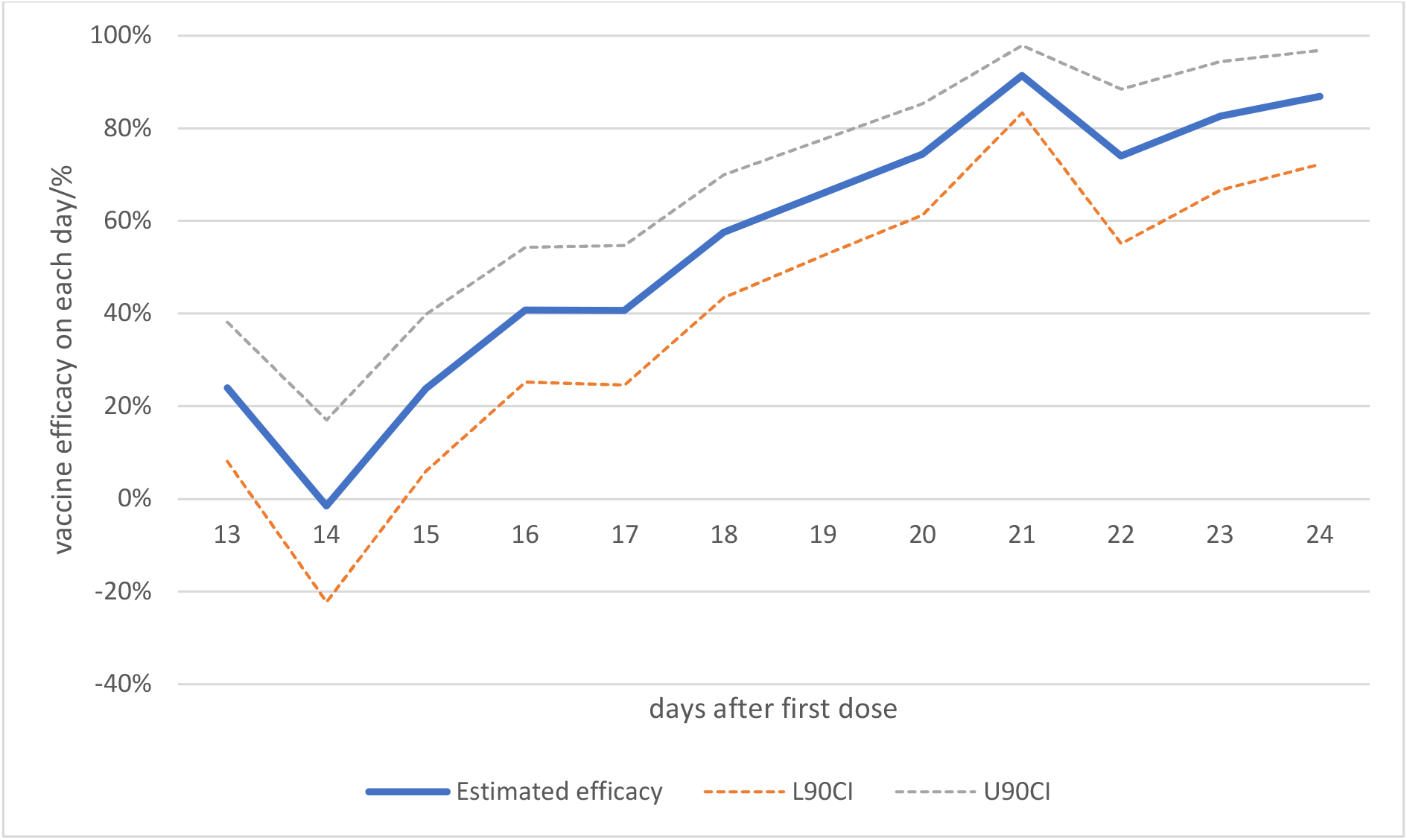
Estimated vaccine effectiveness on each day from day 13 to 24 after a single dose with upper and lower 90% credible intervals.

## Discussion

Our analysis would suggest that the effectiveness of the Pfizer vaccine at least in this Israeli cohort increased gradually day by day from about day 14 till reaching a peak of around 90% effectiveness on day 21 even before any second injection. But there was a strong increase in incidence over the first week after the injection. If the increase in incidence during the first few days after immunization is a result of people being less careful after they have had their first injection, then the vaccine effectiveness after a single dose may be even greater. This analysis would suggest that a single dose of the Pfizer vaccine is able to deliver very high protection, albeit only from about three weeks after the initial injection.

There are a number of weaknesses in the analyses undertaken by the authors of the Chodick et al paper that need to be addressed. Firstly, the reported effectiveness included data from days (13 to 17) when it was apparent that the vaccine was not yet providing much protection and therefore their analyses could not provide any real estimate of the overall effectiveness of a single dose when the second dose was delayed for up to 12 weeks. By analysing effectiveness for each day, the analysis approach presented here overcomes this weakness.

Secondly, by comparing incidence in a later period with that in an earlier period, Chodick et al effectively used a historical rather than a contemporary control group. Whist such historical comparisons can have value they are known to have serious issues; it has been said that “most historical control groups are compromised for some reason” (Streiner & Norman 1998; Baker & Lindeman 2001). Indeed, in one of our own systematic reviews of dengue control strategies those studies using historical controls gave much greater apparent effectiveness (Al-Muhandis & Hunter 2011). In the context of this study, a key issue is whether individuals after their first dose might start to be less careful about social distancing and so increase their exposure risk (SPI-B 2020). Whilst it is too early to have empirical data on this, the finding that incidence increased for a week after first injection could be an indication that such increased exposure does happen. In any event this type of bias would reduce the apparent effectiveness of the vaccine rather than increase it.

Thirdly, the case definition used by the Chodick et al. was less strict than that used in the phase 3 trials and as such were likely to include more cases of mild illness. We know that for most of the available phase 3 trial results for all of the vaccines, that vaccine effectiveness seems to be better at reducing severe disease that at reducing mild disease (Polack et al. 2020; Voysey et al. 2020; Baden et al. 2020). As such we may expect the actual effectiveness of a single dose to be even better for preventing severe disease, hospitalisation and deaths than estimated here.

Whilst we do not know how long this immunity will last beyond 21 days without a second booster, we are unlikely to see any major decline during the following nine weeks. Immunity levels to natural infection with SARS-CoV-2 declines after infection but IgG levels are relatively stable over 6 months and T cell immunity declines with a half-life of 3 to 5 months (Dan et al. 2021).

This analysis of “real world data”, rather than cast doubt on the protective effect of a single injection as the authors of the original study seem to imply, actually provides strong evidence that a single dose of the Pfizer vaccine can provide strong protection from 21 days after the initial injection. As such, the data from this Israeli paper provide strong support for the UKs policy of delaying the second dose of all vaccines.

## Data Availability

All data was extracted from a pre-print already posted on MedRxiv and cited in the paper.

## Notes

### Competing Interest Statement

The authors have declared no competing interest.

### Author Declarations

Not necessary as analyses based only on publicly available summary data

